# Rapid response to haemorrhagic fever emergences in Guinea: toward a community-based system to enhance commitment and sustainability

**DOI:** 10.1101/2025.03.04.25323367

**Authors:** Saa André Tolno, Séverine Thys, Alpha-Kabinet Keita, Maxime Tesch, Chloé Bâtie, Véronique Chevalier, Marie-Marie Olive

## Abstract

Since the 2013-2014 Ebola virus disease outbreak, Guinea faced recurrent epidemics of viral haemorrhagic fevers. Although Guinea has learned lessons from these epidemics by improving its disease surveillance and investigation capacities, local authorities and stakeholders, including community, are not sufficiently involved in response of disease emergence. This led to measure poorly adapted to the local context and consequently less understood and engaging by these local stakeholders. However, recent research has shown that community-based response measures have already demonstrated their effectiveness. By using a qualitative participatory research, this study aimed to (1) describe and analyse the health-related signals that alert local stakeholders, (2) describe the outbreak response measures implemented in Forest Guinea at local and central level, and (3) identify the obstacles and levers for implementing responses adapted to the local socio-cultural context. Ultimately this study should help to build an integrated, community-based early warning and response system in Forest Guinea. Local stakeholders are alerted by a variety of signals: sanitary, environmental and socio-political signals. Regarding health signals, the local stakeholders are supposed to follow a flow chart developed at the central level with a top-down approach. However, our interviews showed that local stakeholders poorly understood this official flow chart. Consequently, we developed, with these local stakeholders, a response flow chart based on their perception and experiences. This diagram, co-constructed with local stakeholders, opens the door to the development of a community-based response. We then identified six main obstacle categories from the interviews, among them lack of logistical and financial resources, lack of legitimacy of community-workers and lack of coordination. Based on the obstacles, we have proposed recommendations for developing a response to emerging zoonotic diseases that would enable local stakeholders to better understand their roles and responsibilities and improve their commitment to the outbreak response.

## Introduction

For 10 years, Guinea faced recurrent epidemics of viral haemorrhagic fevers (VHFs). The emergence of VHFs, in particular the 2013-2016 Ebola virus disease (EVD) epidemic, was a health crisis of unprecedented proportions (1,2). In 2021, two new epidemics were declared in the Forest Guinea region, one reemergence of EVD which started in Gouécké (N’Zérékoré prefecture) totalling 16 reported confirmed cases, among which 12 people died, and one of Marburg haemorrhagic fever in Temessadou M’Boket (Guéckédou prefecture) with one death reported (3,4). In addition, recurrent cases of Lassa fever have been regularly reported in all regions of the country since 2017 (5).

Guinea has learned lessons from these epidemics by improving its disease surveillance, investigation and response capacities. Among others, the existence of a “Response Plan to the Ebola Virus Disease Epidemic” developed in 2021 by the Guinean National Health Safety Agency (ANSS) has improved the response to epidemics by (i) strengthening prevention and control measures of the infections, (ii) supplying health facilities and Epidemiological Treatment Centers (CTEpi) with inputs (medicines, disinfectants, personal protective equipment, etc.), (iii) organizing systematic follow-up of case contacts and their care, (iv) vaccinating case contacts and healthcare workers at risk, and (v) strengthening surveillance at entry points and cross-border collaboration (6). In addition, the construction of several reference laboratories in Conakry, the capital city, and local laboratories dedicated to the early detection and confirmation of cases located as close as possible to communities in high-risk areas allowed to quickly investigate the related-health signals (6,7). Despite the existence of this plan, the international and national institutions involved in the management of these health crises are not sufficiently coordinated, leading to duplication and the implementation of similar tasks during epidemics (8).

VHFs are considered to be zoonotic diseases, with the wildlife being reservoir of some of these diseases. The transmissions of the pathogens of concern at the human-animal interface primarily affect rural communities, which are in frequent contact with domestic animals and wildlife and thus at the frontline of VHFs emergence. VHFs emergence in forest regions is also shaped by the interaction of climate, socio-economic and ecological dynamics. These dynamics are non-linear, and therefore make the emergence of VHFs such as EVD complex and difficult to predict (9). While it is mostly local people who are very familiar with and knowledgeable about the changes in the forest and its fauna, emergence studies have unfortunately not always capitalized this local expertise (10). In addition, local authorities and stakeholders are not sufficiently involved in surveillance and response leading to response activities poorly adapted to the local context (11). This poor adequation and lack of trust between local communities, officials and medical professionals could give rise to protests, occasionally violent, and failure to comply with some response measures. As example, during 2014-2016 Ebola epidemic, secure burials, vaccination of contact cases and people at risk, declaration of community cases were poorly accepted by affected communities (12,13). Community and local stakeholders’ engagement are now commonly regarded as a crucial entry point for gaining access and securing trust during humanitarian emergencies (14–17). However, Le Marcis et al. (2019) emphasized the importance to recognize that within communities, power and legitimacy are always contested resources, and therefore for community engagement tactics during emergencies, no ‘one size fits all’, nor inflexible or top-down responses are appropriate (18). Community-based response measures have already demonstrated their effectiveness, such as during the 2018-2020 Ebola epidemic in the Democratic Republic of Congo where a community-based contact isolation strategy was implemented (15).

In 2019, a study conducted in Guinea by Guenin et al. (2022) exploring community’s capacity to detect emerging zoonoses and surveillance network opportunities showed that the response to an emergence first relies on the surveillance system in place. Its ability to detect abnormal animal or human health events at the community level demonstrated that the capacity for early detection and rapid response to emergencies is based on the diversity of local knowledge of existing diseases, and on the perception of clinical signs. The same survey also showed that local authorities, local agents, and communities were not sufficiently involved in drawing up intervention plans, surveillance and response (19).

The bottom-up approach of co-constructing alert responses to zoonotic emergences, involving different surveillance stakeholders, seems therefore to be an alternative way of helping them to commit to and take ownership of the system (20). Once a health-related signal has been issued, the success of the response depends on the type of alert and the degree to which stakeholders adhere to the response plan (21,22). Consequently, a better understanding and consideration of the priorities, constraints, and levers of local and national stakeholders is needed to adapt the response system, improve its acceptability by stakeholders and finally to improve the system’s ability to rapidly detect and control any emergence.

Qualitative and participatory approaches are effective tools to address complex health issues by considering individual characteristics and societal influence on health determinants. In particular, these methods contribute to increase researchers’ and decision makers’ abilities to consider and understand the complexity of stakeholders’ behaviour (23). They are increasingly used by interdisciplinary teams to enhance stakeholders’ involvement from various sectors, all imbedded in a particular socio-cultural context (23). Among participatory approaches, participatory epidemiology is widely used to improve human and animal disease surveillance (24,25). It is based on the collection of qualitative and semi-quantitative epidemiological data in communities through interviews and visual tools among other methods (26,27). Knowledge and experiences of relevant stakeholders are shared with the research team, leading to the stakeholder’s involvement. Its flexible and stimulating corpus of methods enable to develop intervention and monitoring strategies tailored to the communities involved, considering their socio-economic and cultural constraints (25). Qualitative research has also been successfully used to provide baseline information and identify strategy to develop community-based response to Ebola in Liberia (14).

By using a qualitative participatory research, this study aimed to (1) describe and analyse the health-related signals that alert local stakeholders, (2) describe the outbreak response measures implemented at local and central level in Forest Guinea, and (3) identify the obstacles and levers for implementing plans adapted to the local socio-cultural context and the needs of the stakeholders involved in the response. These specific objectives should ultimately help building an integrated, community-based early warning and response system in Forest Guinea.

## Materials and methods

### Study sites

The study areas included sites at local level in the Guinea forest region in south-eastern Guinea (prefectures of Gueckedou and N’Zerekore) and at central level in the capital city Conakry (Fig 1). Within Guinea forest region, we selected six sites. Four sites were selected in Gueckedou prefecture: Guéckédou town, Koundou sub-prefecture, Temessadou Djigbo sub-prefecture and Temessadou M’Boket village (situated in Temessadou Djigbo sub-prefecture). Two localities were selected in N’Zérékoré prefecture: N’Zérékoré town and Gouécké village (situated in Gouécké sub-prefecture). These sites were chosen because they had been affected by the Ebola epidemic in 2014 and 2021, the 2021 Marburg epidemic or recurrent Lassa fever outbreak (28–30).

**Figure 1:**
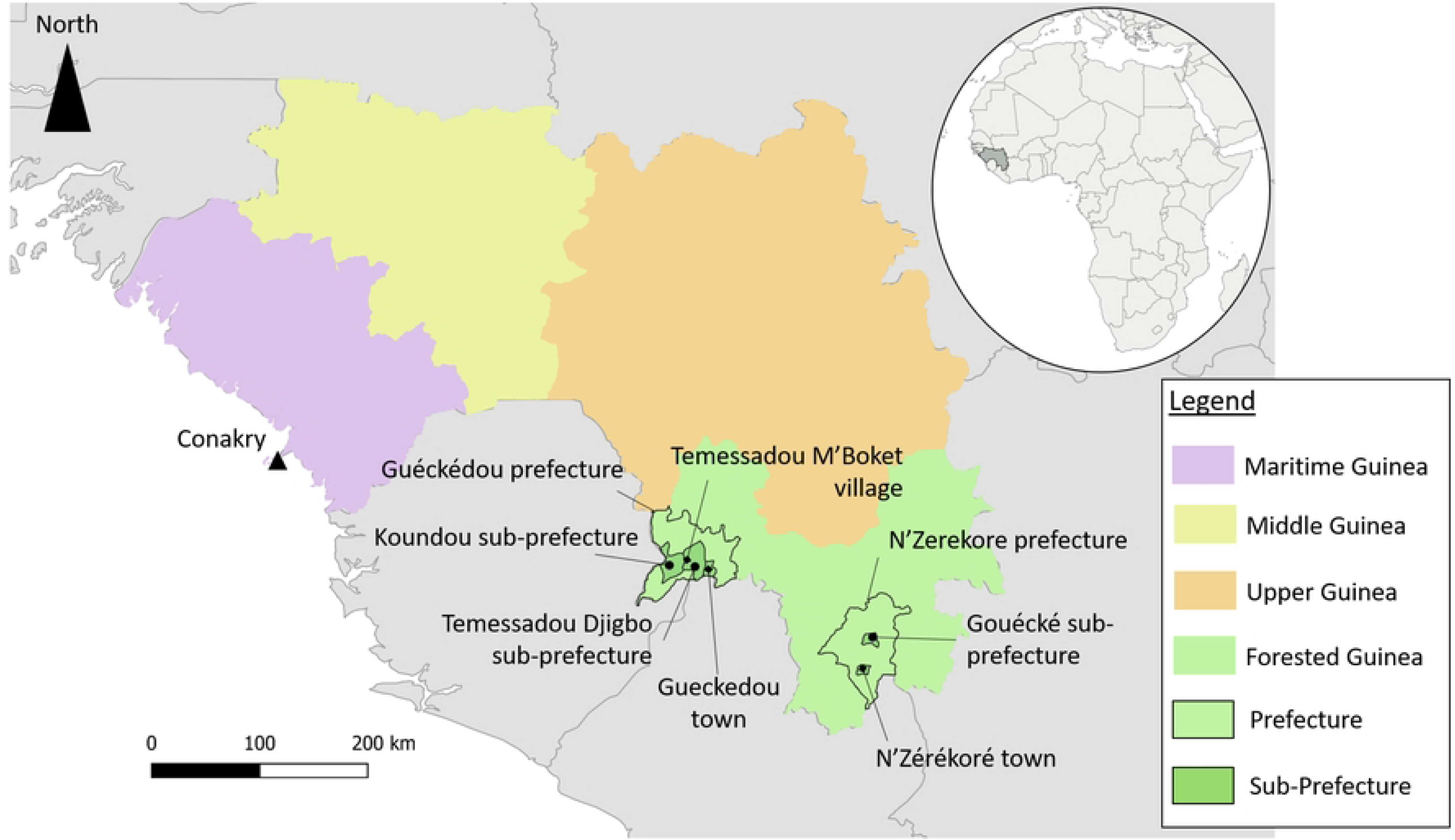
Map of the study sites

### Study design

Firstly, by using bibliography and official documents, we produced a list of stakeholders and administrative structures involved in the surveillance and response to zoonotic disease outbreaks in Forest Guinea (8,19,31). Additional stakeholders were then selected and/or introduced into the study using the snowball sampling method (32).

Based on the theoretical saturation of information and depending on the availability of targeted stakeholders (33), 13 focus group discussions (FGDs),13 individual in-depth interviews (IDIs) and informal discussions were carried out, involving a total of 158 participants, including 124 men and 34 women (Table 1).

**Table 1:** Characteristics of the groups involved in focus group discussions (FGD) or individual in-depth interviews (IDI)

| Type | Category of actors | Data collection sites |  |  |  |  |  |  |  |  |  |  |  |  |  | Phases |
| --- | --- | --- | --- | --- | --- | --- | --- | --- | --- | --- | --- | --- | --- | --- | --- | --- |
|  |  | Temessadou |  | Koundou |  | Temessadou |  | Gueckedou |  | N’zerekore |  | Gouecke |  | Conakry |  |  |
|  |  | Djigbo |  |  |  | M’Boket |  | town |  | town |  |  |  |  |  |  |
|  |  | M | F | M | F | M | F | M | F | M | F | M | F | M | F |  |
| IDI 1 | Prefectural technical service agent - Livestock | - | - | - | - | - | - | - | - | 1 | - | - | - | - | - | I |
| ID 2 | Prefectural technical service agent - Livestock | - | - | - | - | - | - | 1 | - | - | 1 | - | - | - | - |  |
| IDI 3 | Prefectural technical service agent - Environment | - | - | - | - | - | - | 1 | - | 1 | - |  | - | - | - |  |
| IDI 4 | Prefectural technical service agent - Human Health | - | - | - | - | - | - | 2 | - | 1 | - | - | - | - | - |  |
| IDI 5 | Sub-prefectural technical agent - Livestock | 1 | - | 1 | - | - | - | - | - | - | - | 1 | - | - | - |  |
| IDI 6 | Sub-prefectural technical agent - Environment | 1 | - | 1 | - | - | - | - | - | - | - | 1 |  | - | - |  |
| IDI 7 | Sub-prefectural technical<br>agent - Livestock | - | - | - | - | - | - | - | - | - | - | - | 1 | - | - |  |
| FGD 1 | CWs - Livestock | 5 | 1 | 5 | 1 | - | - | - | - | - | - | 6 | - | - | - |  |
| FGD 2 | CWs - Environment | - | - | 4 | - | - | - | - | - | - | - | 7 | - | - | - |  |
| IDI 8 | Sub-prefectural technical<br>agent -Human Health | - | 1 | 1 | - | - | - | - | - | - | - | 1 | - | - | - |  |
| FGD 3 | CWs - Human Health | 7 | 1 | 8 | 3 | - | - | - | - | - | - | 6 | 2 | - | - |  |
| FGD 4 | Community members | 3 | - | - | - | - | - | - | - | - | - | - | - | - | - |  |
| FGD 5 | Community member | - | 4 | - | - | - | - | - | - | - | - | - | - | - | - |  |
| FGD 6 | Community member | - | - | - | - | 9 | 1 | - |  | - | - | - | - | - | - |  |
| IDI 9 | Central service agent - Human<br>Health | - | - | - | - | - | - | - | - | - | - | - | - | 1 | - | II |
| IDI 10 | Central service agent -<br>Livestock | - | - | - | - | - | - | - | - | - | - | - | - | 1 | - |  |
| IDI 11 | Central service agent – One- | - | - | - | - | - | - | - | - | - | - | - | - | 1 | - |  |
|  | Health Committee |  |  |  |  |  |  |  |  |  |  |  |  |  |  |  |
| FGD 7 | Community members | - | 9 | - | 4 | - | - | - | - | - | - | - | - | - | - | III |
| FGD 8 | Community members | 12 |  | 7 | - | - | - | - | - | - | - | - | - | - | - |  |
| FGD 9 | CWs - Human Health | 5 | 2 | 5 | 1 | - | - | - | - | - | - | - | - | - | - |  |
| FGD 10 | CWs - Livestock and CWs_Environment | 6 | 1 | 6 | - | - | - | - | - | - | - | - | - | - | - |  |
| FGD 11 | Sub-prefectural technical agents (Human Health, Livestock and Environment) | 2 | 1 | 3 | - | - | - | - | - | - | - | - | - | - | - |  |
| **TOTAL** |  | 42 | 20 | 41 | 9 | 9 | 1 | 4 | 0 | 3 | 1 | 22 | 3 | 3 | 0 |  |

**Legend** : I = First phase of data collection in Forest Guinea, sub-prefectures of Temessadou-Djigbo, Koundou and Gouécké ; II = Second phase of data collection in Conakry; III = Third phase of data collection, feedback and validation of preliminary results in Temessadou-Djigbo and Koundou sub-prefectures; IDI= Individual in Depth Interview; FGD= Focus Discussion Group; CWs= community workers; - = None.

In the framework of our study, we define: (i) an event as something that may occur in the community and may have a negative impact for the community (34), (ii) a signal as an immediate alert at the early stages of the event (disease outbreak or disaster) which requires an immediate notification and investigation for verification (https://www.emro.who.int/health-topics/ewarn/index.html) and (iii) a response, as actions triggered to stop or limit the consequences of the event. In the context of infectious disease, a response is a set of actions “triggered to stop the spread of an infectious disease swiftly, keeping as few people as possible from being infected” (https://www.taskforce.org/outbreak-response/). In line with these definitions, we defined the process leading to a response as follows: it begins with the detection of a *signal* that alerts local stakeholders that an *event* may occur in this population, requiring actions in *response*.

### Data collection

Field work was carried out between April 2022 and April 2023 at the above-mentioned sites during three separate phases. The first two phases aimed at collecting data while the last phase was a validation step. The first phase took place between April and June 2022 included a total of 91 local participants, in the prefectures of Gueckedou (prefectural services in Guéckédou town and sub-prefectural, community-workers and community members in Temessadou Djigbo, Temessadou M’Boket and Koundou) and N’Zérékoré (prefectural services in N’Zérékoré town and sub-prefectural, community-workers and community members in Gouécké; Phase I, Table 1). The second phase was carried out in March 2023 at central level in Conakry and included 3 participants (Phase II, Table 1). The third phase was organised in April 2023 and included 64 local participants from phase I in Temessadou-Djigbo and Koundou sub-prefectures (phase III, Table 1). During this latter phase, the preliminary results were presented to the participants involved in the first two phases at the study sites in order to clarify, amend and supplement the results presented on the basis of their previous statements and concerns, and to obtain their approval.

The thematic guide was pre-tested during 2 FGDs including 12 participants, and the interview guide was pre-tested with 2 IDIs of stakeholders, all being from the deconcentrated technical services and from the community. After the pre-test phase, a total of 11 FGDs and 11 IDIs were conducted. The discussed topics included (i) the alert and response protocols in place during previous epidemics of Ebola, Marburg and Lassa between 2014 and 2021 at the community level, technical agents and central stakeholders; (ii) the constraints and keys for a successful response; (iii) the needs and expectations of the actors involved in the response mechanisms and, (iv) the official alert response organisation chart and its understanding by actors, from the local to national levels. The number of participants per FGDs varied between 6 and 12 people. Their composition was homogeneous according to stakeholder categories, namely community members, community-workers, local agents, prefectural agents and central service agents, but not necessarily in terms of gender. Indeed, based on the knowledge of the local context, gender-related cultural sensitivity does not influence freedom of opinion in the socio-professional sphere. Environmental community-workers have been associated with livestock community-workers in FGDs because in Guinea the surveillance of wildlife diseases is carried out by the veterinary services (35). FGDs and IDIs lasted between 40 and 90 minutes and were conducted either in French, in the local language (Kissi) or in mixed French-Kissi by the principal investigator. The team was completed by an assistant to ensure that the interview runs smoothly, keeping an eye on the effective participation of the stakeholders and the coherence of the interventions for the debriefing. A reporter was responsible for taking notes as well as making audio recordings and taking photographs. The team members had previously been trained in the use of participatory approaches.

### Ethical framework

The study protocol received authorization from the National Ethics Committee for Health Research (CNERS) in Guinea in accordance with official acts N°028 /CNERS/22 of 19 April 2022 and N°050 /CNERS/23 of 05 April 2023. The approval of the local authorities was requested and obtained in each sub-prefecture of interest, after explaining the objectives of the study to the representatives. Before the interviews begin (individual and FGDs), written consent forms were obtained for each participant or, in the case of FGDs, by a designated representative of the stakeholder category concerned.

Respondents were free to participate to the study without any obligation to answer all the questions. The interviews were recorded using a recording device, and notes and photographs were taken when agreed by participants and relevant to the study. The interviews were anonymised during the processing phase.

### Data processing and analysis

The interviews were transcribed in full and translated into French when necessary (with a Kissi-French translation provided by the principal investigator).

Firstly, the transcripts reviewed, as well as notes from informal discussions, field notes and diagram pictures were imported into Nvivo2022 software (Nvivo 14; QSR International). Secondly, the transcripts were classified according to interview type (FGDs and IDIs), stakeholder category data collection site (Gouécké center, Temessadou Djigbo, Koundou, Temessadou M’Boket, Guéckédou, N’Zérékoré and Conakry) and decision-making level (local, deconcentrated and central). The transcripts, pictures and notes were classified and sortd and the relationships and trends in the data were examined. We conducted a thematic analysis where the main themes were identified using a deductive approach based on the objectives of the study. Then, an inductive approach helped to generate new themes and/or sub-themes emerging from FGDs and IDIs (36). Following the iterative process of thematic analysis, the coding tree used to define the themes was finalized after consensus between main authors.

## Results

Firstly, a mapping of the health-related stakeholders at the local (sub-prefectural), prefectural, regional and central levels have been produced (Fig 2). At the local level, community members identified were local elected representatives, matrons, opinion leaders, healers, hunters, farmers, herders, woodcutters, teachers, and town criers. Community-workers for human, animal and environmental health sectors were also present at this local level, as well as decentralized technical services agents for those three sectors from sub-prefectural administration. At the prefectural level, we identified agents of health, livestock and environment, water and forest Prefectural Departments. The same sectors were identified at the regional level. Finally, at the central level, the main bodies in charge of diseases surveillance and responses were the ANSS, the National Directorate of Veterinary Services (DNSV) and Guinean Office of National Parks and Forest Reserves (OGPNRF), all members of the Guinean One Health Platform.

**Figure 2:**
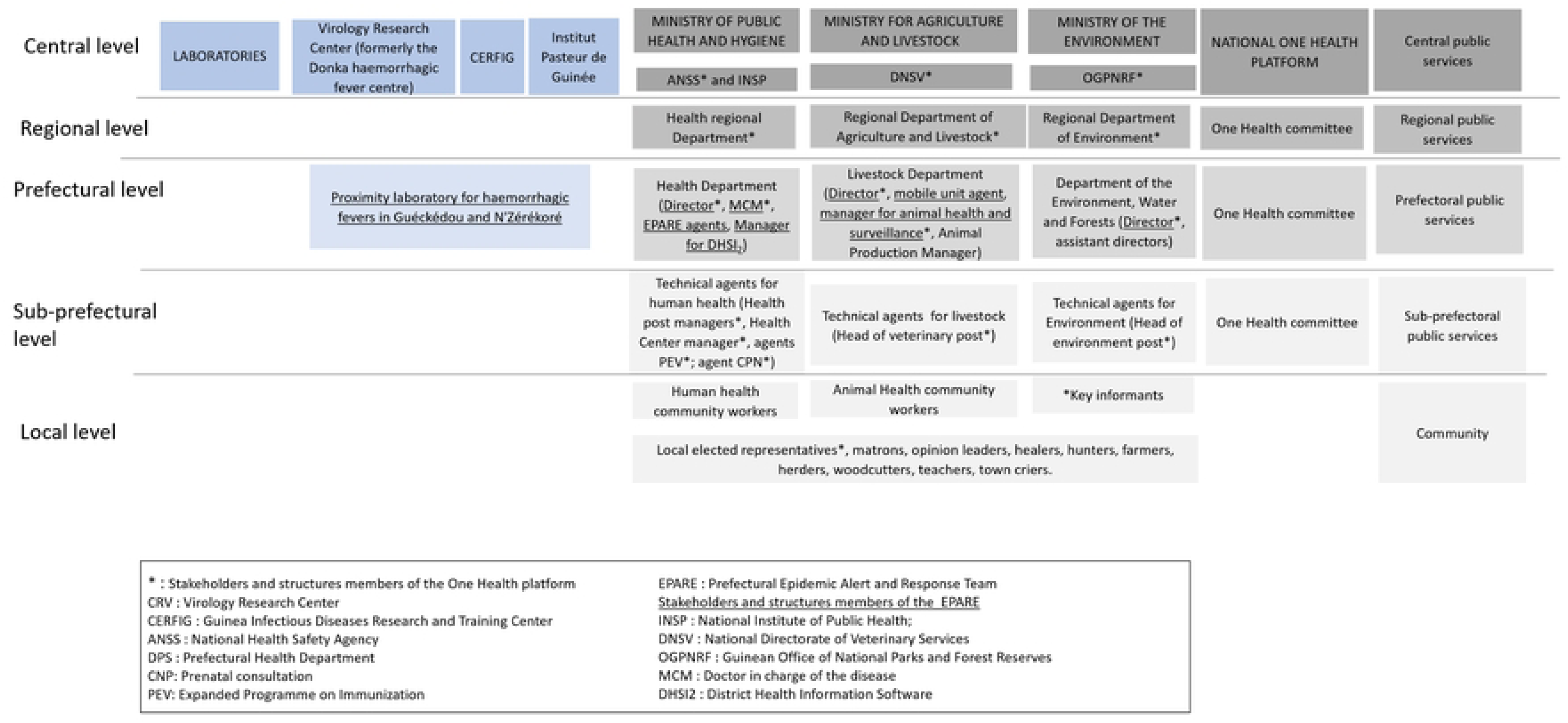
Mapping of stakeholders and structures involved in outbreak response

Among the 158 participants of the study, 131 (82.9%) were community workers and members of the community, who are considered to be front-line actors in the event of alarming events; 24 (15.2%) were technical agents from decentralised services (sub-prefectural and prefectural services), and 3 (1.90%) were central actors. By gender, 34 (21.5%) of the study participants were women, compared with 124 (78.5%) of these participants being men (Table 1).

The results presented below are organised by major theme: (i) alarming events and alerts at local level; (ii) response flowcharts; (iii) obstacles, (iv) levers in implementing response measures and (v) perceptions of control measures. They follow the path that information may take from the alarming event triggering the signal to the implementation of response measures. Any similar and/or different ideas mentioned by the participants are mentioned, considering the category of stakeholder, the data collection site and the level of decision.

### Events and signal detection and notification

Various signals described as alarming or worrying were mentioned by the participants. They were grouped into three categories: health-related signals (human and domestic or wild animal diseases or deaths), environmental signals (bush fires, floods, tornadoes), and socio-political signals (deaths and damages caused by strikes and demonstrations (Fig 3). According to the participants each of these signals could lead to event that may impact either human and/or animal health, environment, well-being, or food security. They alarmed communities because of their impact on daily life, including loss of life, loss of livestock and wildlife, reduced trade and commercial flows, and migration of citizens for fear of tougher response measures in the context of health crises. Regarding health signals, those identified by the majority of participants in the study lead to the production of alert to local and national surveillance authorities. These signals were based on cases that were observable through human and animal mortality and most often unusual clinical symptoms.

**Figure 3:**
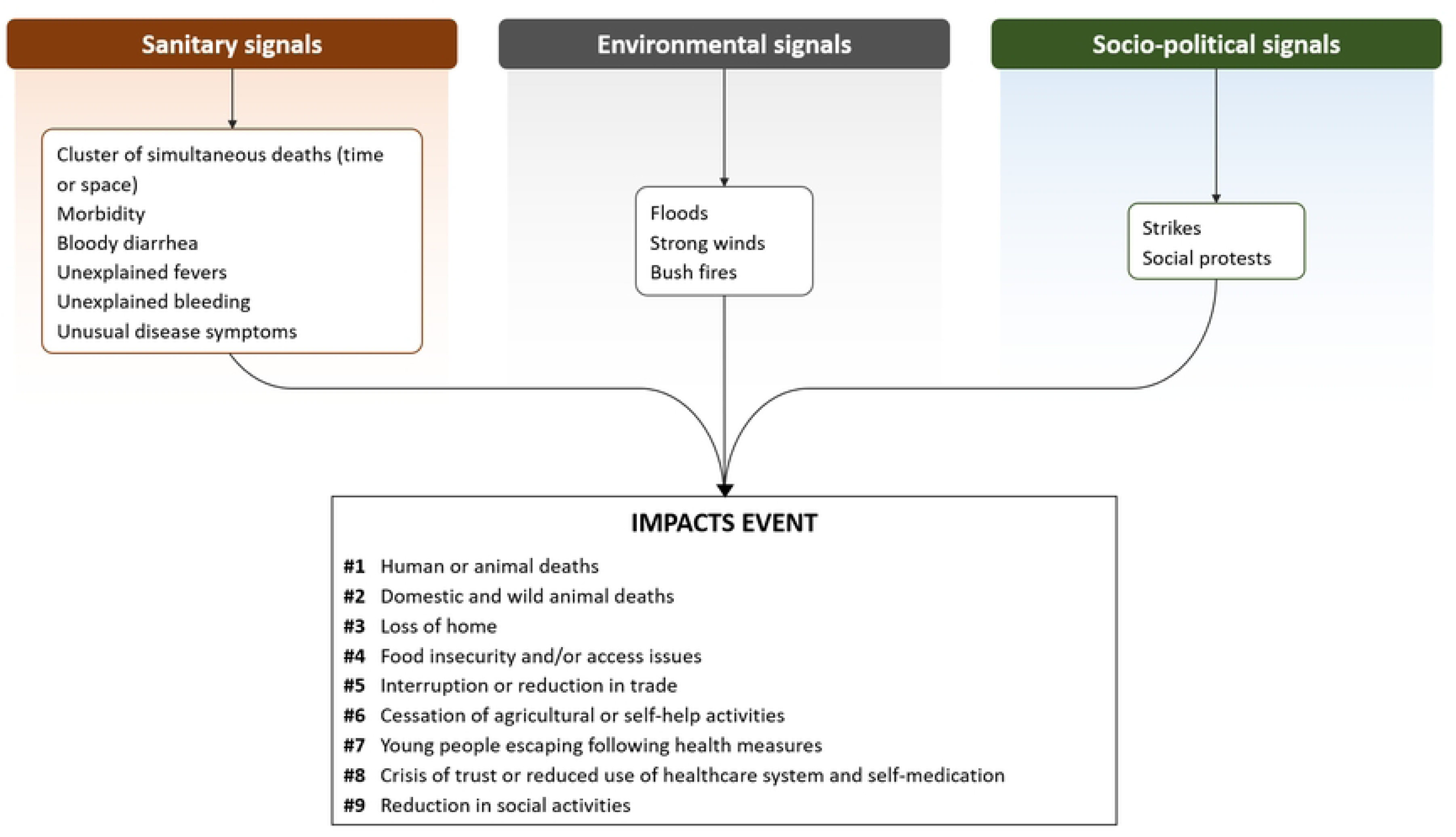
Schematic representation of signals identified by local stakeholders and related negative impacts from FGDs and IDIs conducted in Guinea from 2022 to 2023 ranked from 1 (most impactful) to 9 (least impactful).

Various surveillance stakeholders at local and central level said that signals were produced by people who were recognised by local elected representatives, and who had voluntarily become involved in the communities, so called community-workers, either in human health, livestock and environmental sectors. They also reported that signals were sometimes produced directly by the community members themselves, without any intermediary community-workers. Looking at the discussion on signals as a whole, each category of stakeholder most often alerted on signals that concerned their area of activity. Signals concerning diseases were most often produced by the human and livestock health community-workers, while the environmental community-workers dealt with forestry problems, bush fires and wildlife mortality.

### Response organisation charts

An official public health emergency management flowchart has been developed and has existed in Guinea since the 2013-2014 Ebola virus disease epidemic crises aiming at managing the alert and setting up the response in the event of a disease. This flowchart has been shared by prefectural epidemic alert and response team (EPARE) at N’Zérékoré site, and schematized in the figure 4. Supervised by the EPARE, this response organisation flowchart defines the steps to be followed from the signal. This plan includes several transitional stages between the signal, the alert and the implementation of the response to a health event, corresponding to the steps required to verify and validate the health event by various entities, until the corresponding response action is implemented. Participants at the local level were asked about their understanding of this flowchart. According to the technical services employees interviewed, this flowchart was difficult to understand and use, as it contained unconventional abbreviations and acronyms, as well as a lack of colour legend and meaning. The official chart identified the emergency operations centre (COU-SP) as the main body responsible for response activities in the ANSS and did not clearly indicate the role and tasks of the other entities involved in implementing the response, including local actors. The level of activation of the response mechanisms in the flowchart depended on the alert threshold and epidemiological threshold for activating the response at the level of the COU-SP and the EPARE. However, the threshold, the timeframe for these actions, and the people responsible for them at the local level were not specified.

**Figure 4:**
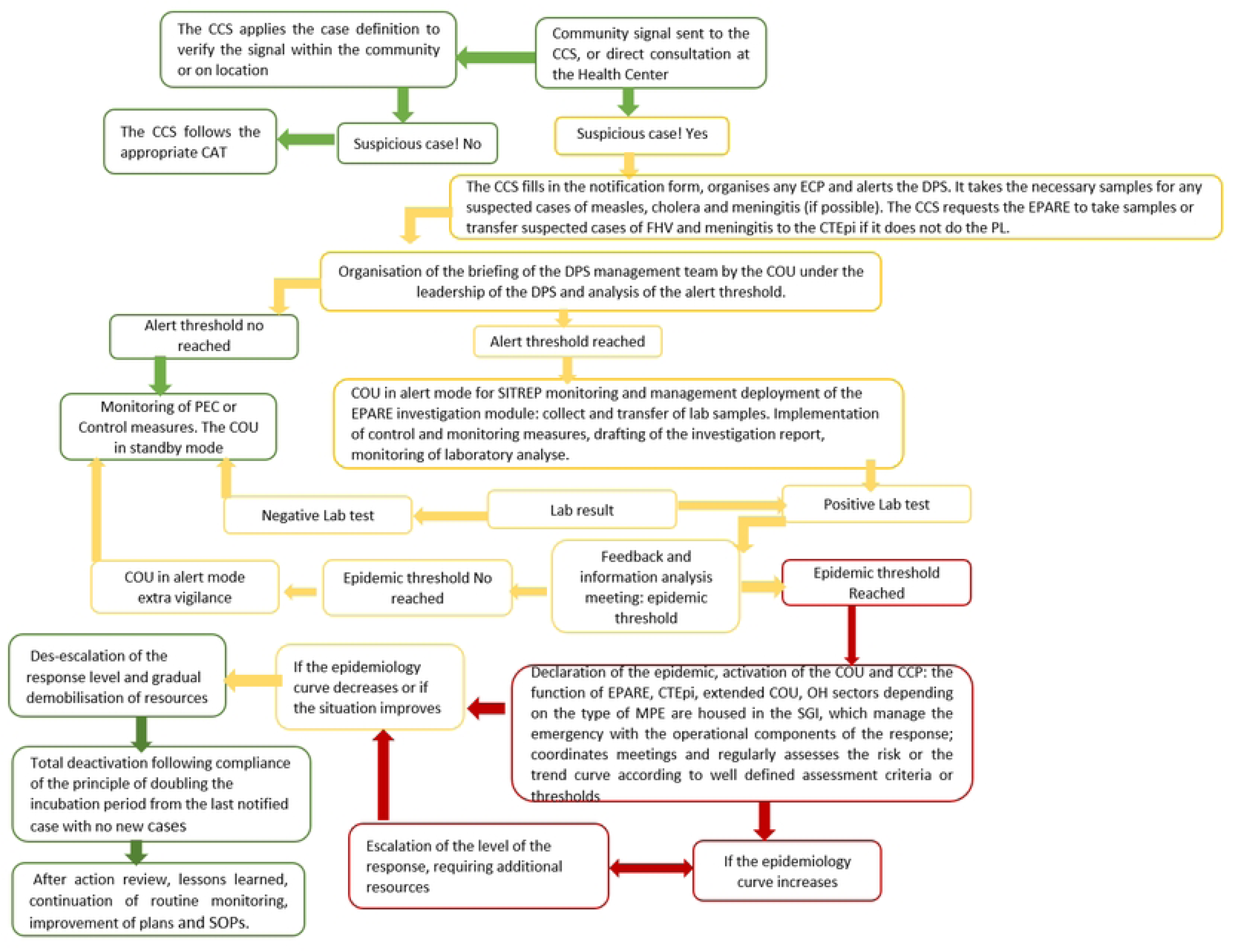
Schematized public health emergency management flowchart (Source: EPARE-N’Zerekore). The acronym legend was produced by the authors on the basis of interviews. CAT: How to proceed; CCS: Head of health centre; CTEPI: Epidemiological Treatment Centre; COU: Emergency Operations Centre; DPS: Prefectural Health Department; EPARE: Prefectural Epidemic Alert and Response Team; SITREP: Situation Report, Lab: Laboratory; MPE: Not determined SGI: Incident Management System; PEC: Care; PL: Not determined

Although this official flowchart was difficult to understand for community-workers, they still took actions based on their understanding of this flowchart and on past experience with response to epidemics. This perceived flow chart was reconstituted by the research team based on the information provided during IDIs and FGDs sessions (Fig 5). During the phase III, this reconstructed flow chart was then amended and validated by the same agents at local level (Fig 5). The perceived organisational chart described the roles and activities carried out by local agents as part of the response in relation to the communities. This flowchart comprised several stages, from the signal to the implementation of the response at local level by the community-workers and local agents and then at the central level. The difference between the official and perceived organisation charts lied in the description and distribution of tasks among all the stakeholders. In our reconstructed organization chart, the alert starts from community members or community-workers. This signal is first verified by sub-prefectural deconcentrated technical agents from decentralized services, under the supervision of the EPARE. If the signal is positive, samples are sent to the prefectural laboratories and/or the local VHFs laboratory (Fig. 2 and Fig. 5). Then, in terms of response, each entity (community-workers, services), and agents of central services respond in different ways if the laboratory results are positive or negative. If the result is negative, all stakeholders provide feedback to the other agents. Especially, community-workers and technical agents from deconcentrated services provide feedback to community. If the result is positive, awareness-raising actions, crisis meetings, training and direct responses such as contact follow-up, transfer of patient, sanitary zoning, are organised by community-workers, sub-prefectural technical agents under the supervision of EPARE. This flow chart show that community-workers play a key role in implementing local response measures. They alerted neighbours and local elected officials. They also raised awareness among the population in order to promote and encourage basic attitudes of self-protection and control. For example, in the event of a suspected zoonosis, community-workers were advised to self-report any suspicions, adopt good hand-washing practices, social distancing, and prohibit handling and eating animals considered to be reservoirs of pathogens, as well as the corpses of other animals.

**Figure 5:**
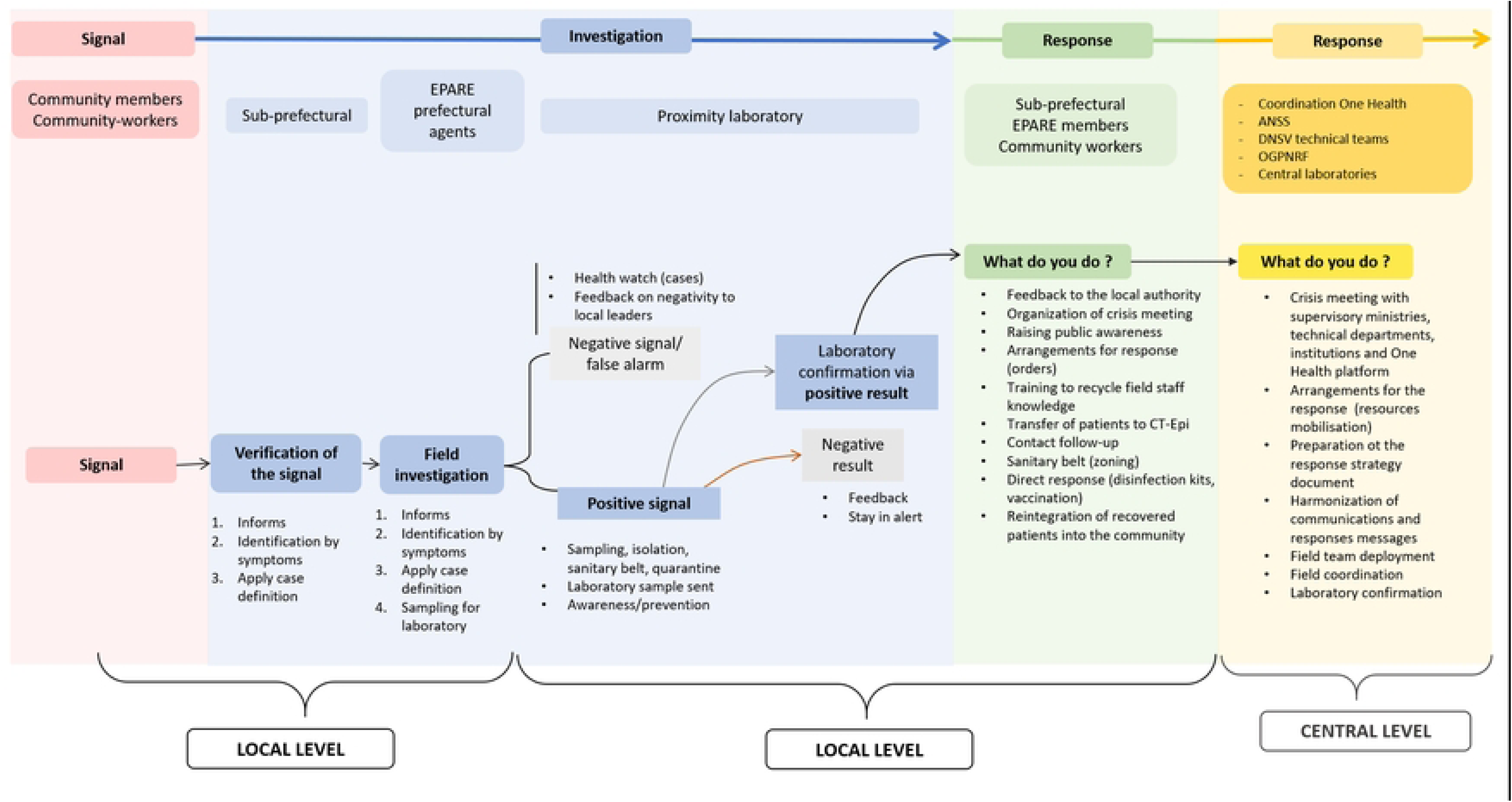
Reconstruction of the perceived response in the case of infectious disease signal. ANSS: National Health Safety Agency, DNSV: National Directorate of Veterinary Services, EPARE: Prefectural Epidemic Alert and Response Team, OGPNRF: Guinea Office of National Parks and Forest Reserves.

The IDIs carried out with the central stakeholders (livestock services, national One Health and human health platform) showed that the response was generally implemented following verification of the signal at the local level by the decentralised technical stakeholders and confirmation of the samples by laboratories.

### Obstacles and levers to implement effective and sustainable response measures

Six main obstacles categories have been identified from the interviews: (1) lack of logistical and financial resources, (2) lack of technical skills in surveillance and early detection, (3) lack of legitimacy, (4) lack of coordination, (5) the large number of actors from health institution in the villages leading to (6) communities’ weariness in regarding to the multiple and uncoordinated actions. According to a prefectural livestock officers, an organisational obstacle lived in the disparity in knowledge between the different players in the One Health sector. He highlighted that human health stakeholders benefited from more training and financial resources than other sectors. Similarly, livestock and the environment local agents stated that the financial, logistical and technical capacities of the Ministry of Health were greater than those of the two other ministries (i.e. livestock and the environment). This would lead to a possible reduced involvement and motivation in these two sectors due to insufficient resources (financial means) for the regular monitoring of post-impact activities and ongoing awareness-raising.

One other category mentioned was the lack of technical skills on surveillance and response to zoonotic disease emergences. According to the agents of the central livestock services, the Ministries of Health and Public Hygiene and the Ministry of Livestock (National Veterinary Services Department) agents have basic skills related to their profession, but there is a lack of initial training in surveillance and response when they integrate their new position. Similarly, in the Ministry of the Environment, although a wildlife disease surveillance system is currently being developed, they still suffer from a shortage of competent wildlife disease surveillance officers. It was reported by field workers as an obstacle to the early detection of wildlife diseases and the effective flow of health information.

The role of the community-workers is not always formalised, which makes their work difficult and lead to a lack of legitimacy.

In addition, there can be inconsistencies and duplication of response activities between international institutions, which is seen as an obstacle by the communities of Temessadou and M’Boket. This lack of coordination between local authorities, research institutes and international institutions during outbreaks was reported by the participants. They highlighted a need of coordination during outbreaks especially for the benefit of communities. As example, during Ebola or Marburg outbreaks, some of the research and response activities, such as bat sampling in caves and human sampling, were duplicated within villages, stifling communities and provoking negative reactions to the presence of repeat teams in villages. Indeed, a community stakeholder reported: *“Coming to ask us every day about the Marburg disease, we are tired, they came here after three months of investigation declare the end of the disease and tell us that we are free to move. After that, we were sent to Guéckédou to the rural radio station to talk about this disease and the means of prevention and control that the village has accepted; so if the disease has been declared defeated, a few months later I see other teams here, people have started to be afraid and to murmur in order to oppose and chase them away. The same things, it’s worrying for us and we need to express our dissatisfaction with certain things” (translated from Kissi, FGD/community/Temessadou M’Boket)*.

These multiple and uncoordinated actions lead to communities’ weariness favouring unacceptance of new actions. Local people in Temessadou M’Boket perceived investigation, responses and research actions as an invasion of their territory, hindering the implementation and acceptance of response measures, as expressed in the following verbatim: “*False promises are sometimes incorporated into communications from institutions or technical partners, leading to subsequent claims by communities.” (translated from Kissi, FGD/community/Temessadou M’Boket)*

From these obstacles, three main categories of needs, have been identified: (1) financial, (2) logistical and (3) trainings.

Human health, livestock and environment community-workers and sub prefectural agents mentioned during the FGDs the need to be granted with lump sums. Additionally, to the formalisation of their status, this would represent a significant boost to their involvement in surveillance activities and the implementation of response measures. Furthermore, financial resources to run health structures (village watch committees, village One Health platforms), telephone credits and fuel costs to facilitate travel to certain remote and isolated areas were also listed as key needs. Closely linked to financial lever, personal protective equipment such as gloves, boots and coats for the rainy season were identified as need to implement measure responses properly. Means of transport, awareness-raising sheets and picture boxes to help understand the diseases and associated signs were also requested. All the FGDs and IDIs at all levels and collection sites emphasised the need for training such as wildlife disease surveillance and sampling procedure, for local and prefectural agents of the deconcentrated technical services. For community-workers (human health, animal, environment), training in disease recognition, prevention and control methods, as well as risk awareness and communication were formulated. In addition, it was recognised by the same actors that reviewing and refreshing the knowledge acquired during previous training courses was essential. The revitalization or installation of health monitoring structures and One-Health platforms was mentioned as a necessity or need.

## Discussion

Lessons learned from several outbreaks show that local community and local stakeholders’ engagements are crucial for effective outbreak responses (14,16,17). Our qualitative study enabled us to give a voice to these local actors to better understand the health-related signals that alert them and describe the outbreak response measures implemented at local level. In addition, our study allows to identify the obstacles and levers for implementing community-based response plans adapted to the socio-cultural context and the needs of local stakeholders in Guinea.

### Main findings

Several alarming signals, ie. sanitary, environmental and socio-political, have been identified by local stakeholders. These signals have led to events that may impact either human and/or animal health, environment, well-being, or food security. Regarding sanitary signals, we described the official outbreak response flow chart and the one perceived by local stakeholders. The first one has been developed at the central level with a top-down approach whereas the second one has been developed, within this study, with local stakeholders with a bottom-up approach.

The first diagram flow seems to be well adapted to central and decentralized stakeholders who need an overview of the sequence of actions and actors required to respond to outbreak. Conversely, since it contains acronyms and terms that are not explained on the document, such as alert threshold, it is difficult to be understood by local stakeholders. Both flow charts start with a signal produced by the community or community-workers. However, the official diagram seems to focus on signals produced in human health, whereas the diagram we have produced is adapted to all three sectors. Our work highlights that community-workers of the three sectors produce alerts relating to signals specific to their sectors.

According to the various statements made by local and central agents about the success of the response teams, some delays in implementing the response measures were still reported. For example, in some localities, citizens opposed the installation of vaccination tents during Ebola 2021. The same findings or similar results were observed during the previous Ebola epidemics of 2018 in the DRC and West Africa in 2014. Communities were imposed intervention measures compared to the implementation of community-based intervention strategies as it had been the case in Liberia and the DRC and which were better accepted (14;15).

Timeliness, such as early detection of signal and early investigation, have been highlighted as key element for an effective response outbreak (37–39). In addition, the clarity of the role and responsibilities of each stakeholders with good coordination among these various stakeholders at different levels constitutes another key element (17,37). The lack of knowledge about to whom to report was, for example, considered as a weakness for community-based surveillance for the Sudan Disease Virus in 2022 in Uganda leading to delay in disease detection (17). From this study, we can propose concrete health signals and response framework developed by community members and community-workers. By this way, local stakeholders would be aware of their roles and responsibilities in surveillance and response outbreak. Previous researches have demonstrated the essential role of community-based surveillance and response systems in the early detection, investigation and response to outbreaks such as Sudan disease virus in Uganda as well as measles and monkeypox in Cameroon (17,40). These later findings highlight the importance of developing surveillance, investigation and response flow chart understood by community and community-workers. Building these flow diagrams with local stakeholders as we have done, could be an effective way of enabling them to take ownership, and consequently better integrate their roles and responsibilities in responding to health emergencies. Additionally, lack of all-cause mortality surveillance was identified as gaps potentially contributing to delayed outbreak detection (17). Our results show that from a community point of view, signals leading to adverse events such as mortality and/or morbidity, whether infectious or environmental, are just as important. This opens the door to the implementation of an integrated surveillance and response system that would consider health events and natural disasters as signals that could have an impact on public health.

### Using qualitative research to discuss complex health issues and identify levers

Measures decided with a top-down approach, without considering the constraints and obstacles of field stakeholders; have shown that they are not accepted and are therefore ineffective and unsustainable (41,42). The reasons for this lack of community acceptability involve complex processes. Qualitative and participatory approaches were successfully effective to discuss complex health issues by considering individual characteristics and societal influence on health determinants and develop acceptable health management in Uganda (17). Indeed, the mobilisation of qualitative and participatory approaches enabled us to apprehend the local knowledge of populations and to identify upstream key actors and other hidden or hard-to-reach stakeholders (26,32). The inclusion of different categories of stakeholders in our study allowed us to collect a diversity of perception.

### Limits

However, a few biases should be mentioned. Firstly, the memory bias which corresponds to the way in which an event is remembered. It can affect results because of forgetfulness or systematic error in thinking by respondents (43). In our study, the interlocutors only remember recent events, such as 2021 outbreaks of Marburg and Ebola, or cannot accurately describe events that occurred several years ago, such as 2013-2016 Ebola outbreak. However, we considered that it was minimized by the large number and wide range of stakeholders interviewed, the triangulation of the data and the amendment and validation the findings by interviewees (44). Some categories of stakeholders, mostly central representatives, were under-represented because of their unavailability and busy schedules. This has led to produce a perceived respond flow chart only from the local stakeholder’s point of view. We can also note a social bias with the low participation rate of women in the FGDs. It constitutes a bias because women had been identified as the preferred interlocutors for family health issues in a previous study (19). The low participation rate of women in the study can be explained by the fewer number of women working in technical services such as veterinary services at national level. Thus, our results need to be taken with caution as the management of alert and response mechanisms might be different if more women were included in our study. We partially compensate this bias with the participation of community midwives and health centres, who are perfectly familiar with the health problems of women and children in the villages, in the FGDs.

Finally, our study was conducted in Forest Guinea where the main outbreaks occurred. It could be interesting to extend this survey to other non-peri-urban sites and to regions of Guinea that have not experienced major epidemic crises. Having a wider diversity of stakeholders would help adapting alert and response systems to public health emergencies at national level with a view to curbing future epidemics. Our survey should also be disseminated to policy-makers to improve rapid respond measures

### Recommendation for overcoming obstacles

Indeed, several recommendations can be made from this perspective. Firstly, while the official response flow chart appears to be well-suited for the central, regional and prefectural levels, a simplified version could enhance the engagement of local stakeholders in outbreak response. The reconstructed flow chart we proposed, based on local stakeholders’ interviews and validated by them could serve as a useful starting point to co-construct a better suited and integrated response system.

Secondly, throughout our study, participants highlighted the financial, logistical and training disparities between human health sectors and both animal and environmental health sectors. These disparities may lead to weaker involvement and motivation of animal and environmental health stakeholders. Similar findings were observed in the same contexts during the prioritisation of diseases to monitor at national level in Guinea (19) and during the investigation of the EBOV alert system management tool in southern Sudan (46). To address these disparities, sharing local resources - through the local One Health platform, for example - and taking a long-term approach the training and logistical needs of each sector would help equalize and bridge the gaps between the three sectors

The lack of legitimacy and financial resources of community-workers has also been highlighted. Indeed, lack of material and financial incentives, was mentioned by the community-workers and local agents in all the FGDs and IDIs to boost and improve the production of alerts for community-workers. This could lead to a loss of motivation on the part of these players, who are nonetheless essential for relaying information from the source of emergences. Community acceptance of community-workers and motivation of these actors have been identified as drivers of success for community-based surveillance (47). As already observed in Ghana, Sierra Leone and Southern Sudan, the training opportunities, material and financial incentives such has telephone credits, cover of travel costs, can therefore be seen as one of the keys to success in the production and reporting of alerts (46–48).

The overall resilience of healthcare systems of countries partially depends on the ability of these countries to rapidly detect and respond to outbreaks. Community-based response systems may enhance this rapid response, provided the incorporation of the key concepts of epidemiological disease surveillance and consideration of the social dynamics of communities.

## Data Availability

The anonymous transcript of the interviews would be available in a dataverse uppon request to the corresponding author.

## Acknowledgements

We would like to thank the National Department of Veterinary Services (DNSV) and in particular Dr. Zogbelemou, prefectural director of livestock in Macenta, and Mr. Leno, Head of veterinary center in Téméssadou, for their assistance in the field. Thanks to Celia Lacomme, research engineer at CIRAD, for her logistical and organizational support. Thanks to all the prefectural and sub-prefectural authorities for their understanding and welcome. Thanks to all participants and local authorities for their participation and the trust they placed in us.

## Funding

This work was supported by the European Union under the Agreement FOOD/2016/379-660, for the implementation of the Action EBO-SURSY “Capacity building and surveillance for Ebola Virus Disease” (https://rr-africa.oie.int/en/projects/ebo-sursy-en/). Saa André Tolno received a PhD fellowship from the Service de Coopération et d’Action culturelle (SCAC) of the French Embassy in Guinea. He was also supported by AfriCam project, funded by the French Development Agency (AFD) as part of PREACTS (PREZODE in action in the global South) program. The funders had no role in study design, data collection and analysis, decision to publish, or preparation of the manuscript

## Notes

### Competing Interest Statement

The authors have declared no competing interest.

### Funding Statement

Yes

### Author Declarations

The study protocol received authorization from the National Ethics Committee for Health Research (CNERS) in Guinea in accordance with official acts N°028 /CNERS/22 of 19 April 2022 and N°050 /CNERS/23 of 05 April 2023.

